# One year of child neurology telemedicine: a data-driven analysis of 14,820 encounters

**DOI:** 10.1101/2021.12.15.21267868

**Authors:** Michael C. Kaufman, Julie Xian, Peter D. Galer, Shridhar Parthasarathy, Alexander K. Gonzalez, Katherine Helbig, Sarah McKeown, Marisa S. Prelack, Mark P. Fitzgerald, Sansanee Craig, Salvatore C. Rametta, Sara E. Fridinger, Uzma Sharif, Susan E. Melamed, Marissa DiGiovine, Lawrence Fried, Marissa P. Malcolm, Sudha Kilaru Kessler, Madeline Chadehumbe, Christina L. Szperka, John Chuo, Laurel Caffee, Donna J. Stephenson, Brenda L. Banwell, Ethan Goldberg, Nicholas S. Abend, Ingo Helbig

**Author notes:** **Corresponding author** Ingo Helbig, MD, Division of Neurology, Children’s Hospital of Philadelphia, 3401 Civic Center Blvd., Philadelphia, PA 19104.

## Abstract

**Introduction:** Determining the long-term impact of telemedicine in care across the diagnostic and age spectrum of child neurology during the COVID-19 pandemic and with the re-opening of outpatient clinics.

**Methods:** An observational cohort study of 34,837 in-person visits and 14,820 telemedicine outpatient pediatric neurology visits between October 1, 2019 and April 9, 2021. We assessed differences in care across visit types, time-period observed, time between follow-ups, patient portal activation rates and demographic factors.

**Results:** 26,399 patients were observed in this study (median age 11.4 years [interquartile range, 5.5-15.9]; 13,209 male). We observed a higher proportion of telemedicine for epilepsy (ICD10 G40: OR 1.4, 95% CI 1.3-1.5) and a lower proportion for movement disorders (ICD10 G25: OR 0.7, 95% CI 0.6-0.8; ICD10 R25: OR 0.7, 95% CI 0.6-0.9). Infants were more likely to be seen in-person after re-opening clinics than by telemedicine (OR 1.6, 95% CI 1.5-1.8) as were individuals with neuromuscular disorders (OR 0.6, 95% CI 0.6-0.7). Racial and ethnic minority populations and those with highest social vulnerability had lower rates of telemedicine participation throughout the pandemic (OR 0.8, 95% CI 0.8-0.8; OR 0.7, 95% CI 0.7-0.8).

**Discussion:** Telemedicine implementation was followed by continued use even once in-person clinics were available. Pediatric epilepsy care can often be performed using telemedicine while young children and patients with neuromuscular disorders often require in-person assessment. Prominent barriers for socially vulnerable families and racial and ethnic minorities persist.

## Introduction

The COVID-19 pandemic shifted much of non-urgent outpatient care to telemedicine, including care of children with neurological disorders.^1^ However, while several studies had demonstrated the overall feasibility of telemedicine,^2-8^ there was a paucity of data describing the outcomes of patients treated through telemedicine, especially in child neurology.^9-11^ Child neurology assessments often rely on physical exams, so providing adequate care using telemedicine can be challenging, exacerbated by the relatively high proportion of rare conditions.^12-14^ We have previously shown that telemedicine visits in child neurology are adequate for approximately 95% of all visits^15^ but, in parallel with other studies, we assessed telemedicine soon after its implementation. As telemedicine use stabilized during the pandemic, new data became evident, such as differences in telemedicine access among underrepresented racial and ethnic minorities.^16^

We aimed to understand the overall utility of telemedicine beyond its initial urgent implementation. The period in which both in-person and telemedicine visits were available as part of our care paradigm offered the opportunity to assess how providers and families used telemedicine versus in-person care. We studied all outpatient visits within our large child neurology practice to identify trends in telemedicine utilization across demographic, clinical, and social vulnerability variables.

## Methods

### Standard protocol approvals, registrations, and patient consents

Data analysis was completed retrospectively from ongoing care and met exemption criteria per the Institutional Review Board at Children’s Hospital of Philadelphia. Written consent was not required.

### Setting

Our study was performed within a pediatric specialty care network including an urban quaternary care hospital and ambulatory center along with eight satellite locations in Pennsylvania and New Jersey. The practice includes general child neurology as well as specialty programs with ∼32,000 visits per year from local, national and international regions.

### Implementation of telemedicine visits

Telemedicine visits were first implemented in March 2020.^15^ In brief, during the first month of the pandemic, audio-video telemedicine was used for all new patient visits and most established visits. Telephone visits were used for established patients who lacked access to video visits. We used the telemedicine software embedded in the Epic system (Verona, WI) in which patients and caregivers participated by video through a mobile phone application called MyCHOP.

Telemedicine visits required an active client-side patient portal at the time of the encounter, enabling us to track scheduled telemedicine visits that resulted in successful activation or an already active patient portal at the time of visit. For each week, we determined activation rates as a proportion of scheduled encounters through a HIPPA compliant telemedicine platform.

### Phases of telemedicine use

We distinguished between several phases of telemedicine use, including the (1) in-person only “prior phase” before the onset of the pandemic, during which no telemedicine visits were performed, between October 1, 2019 and March 15, 2020, the (2) “shutdown phase” between March 15, 2020 and April 12, 2020 during which telemedicine was performed almost exclusively, the (3) “ramp-up phase” between April 12, 2020 and June 5, 2020 during which in-person visits were re-introduced, and the “steady-state phase” between June 5, 2020 and April 9, 2021 during which the rate of in-person visits to telemedicine visits was relatively stable and in-person visits accounted for 50% or more of all outpatient visits.

### Data extraction

Clinical documentation was performed using a single electronic medical record system accessible via the Epic Clarity database. We identified all outpatient encounters and extracted demographic variables including age, sex, race, and ethnicity, along with geographical regions. This was then mapped to the 2018 Centers for Disease Control and Prevention Social Vulnerability Index (SVI), segmented into ranked quartiles.^17^ We extracted the primary ICD10 code of patient encounters (e.g., G40 instead of G40.10) to stand in for diagnoses. We used our previously developed Natural Language Processing pipeline within Oracle SQL to detect the provider questionnaire within the full-text and parsed the semi-structured answers.^15^ Data are presented as descriptive statistics and compared using Fisher’s exact tests, Wilcoxon Rank Sum tests, and Mann-Kendall tests using the tidyverse package in the R Statistical Framework.^18,19^

### Follow-up within intended time window

We determined the number of individuals who were expected to follow-up based on the intended follow-up period documented from their previous encounter. We reviewed assigned follow-ups since October 2018 to accommodate 12-month follow-up times for the beginning of our data collection and then assessed the proportion of individuals who did not have follow-up visits by this date plus a 30-day grace period. We referred to this ratio as the Care Window Index (CWI), indicating the proportion of individuals who were unable to follow-up or had follow-ups outside of the intended timeframe relative to those within the timeframe. We also categorized certain individuals as “lost to follow-up”, defined as any individual who was supposed to return over the previous year but did not return. This included individuals who had not followed up since March 15, 2020.

## Results

### Patient demographics

Our cohort included 26,399 individuals with 34,837 in-person visits, 14,820 new and follow-up telemedicine visits and 1,554 telephone visits (Table 1). SVI could be determined for 99% of individuals (26,128/26,399), including 43% (11,226/26,128) with lowest SVI, 26% (6,900/26,128) with medium-low SVI, 16% (4,055/26,128) with medium-high SVI, and 15% (3,947/26,128) with highest SVI. Of the individuals with highest SVI, 48% (1,880/3,900) were self-identified as Black and 24% (930/3,909) were self-identified as Hispanic or Latino, indicating that most individuals with highest SVI were underrepresented ethnic and racial minorities. We observed a higher use of telephone visits (odds ratio [OR] 1.7, 95% confidence interval [CI] 1.5-2.0) and a lower use of telemedicine (OR 0.7, 95% CI 0.7-0.8) in individuals with highest SVI, but we did not observe differences between other SVI categories.

**Table 1.**
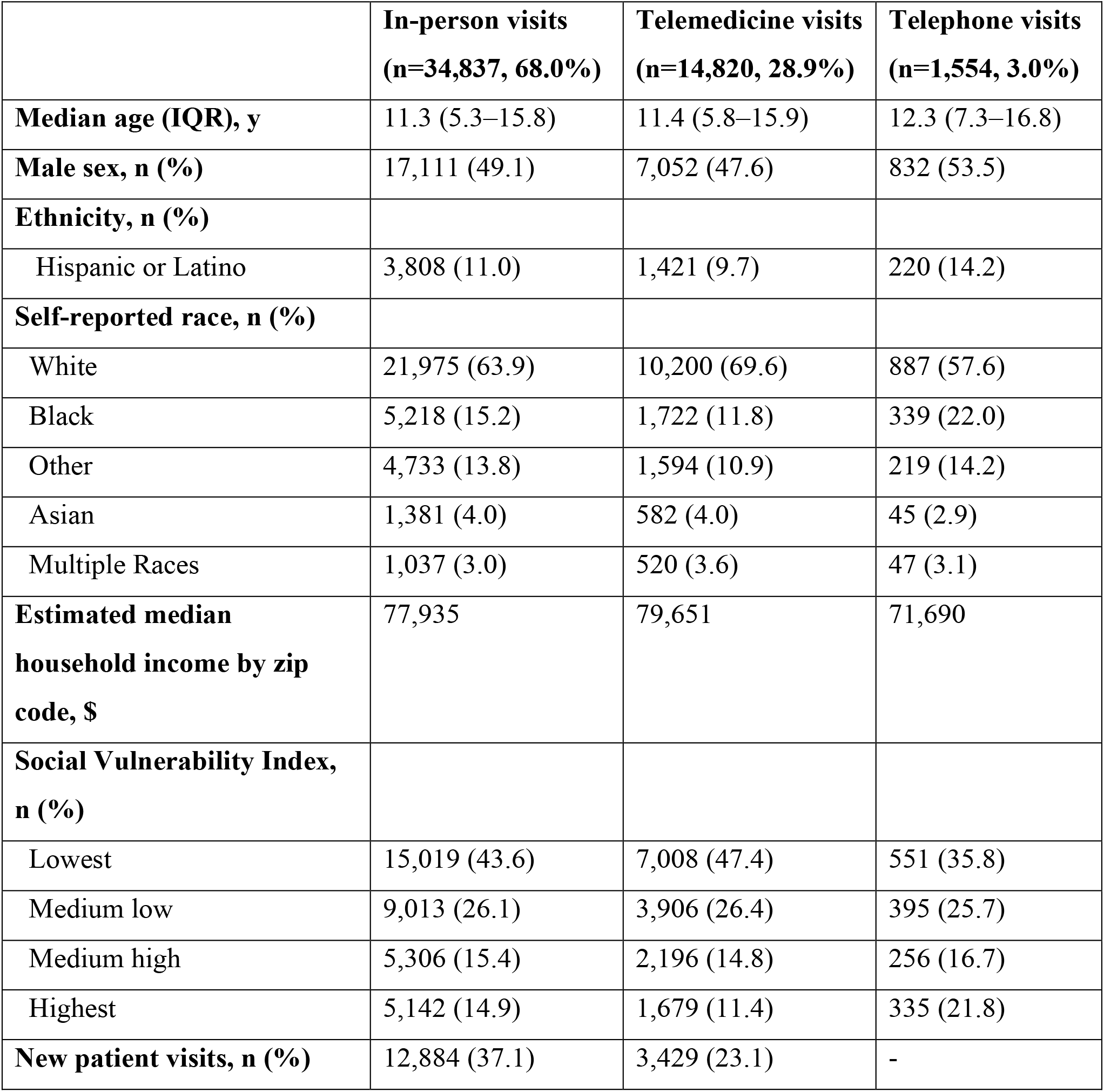
Demographic data of child neurology outpatient visits from October 1, 2019 through April 9, 2021, segmented by in-person, telemedicine and telephone visit types.

|  | **In-person visits**<br>**(n=34,837, 68.0%)** | **Telemedicine visits**<br>**(n=14,820, 28.9%)** | **Telephone visits**<br>**(n=1,554, 3.0%)** |
| --- | --- | --- | --- |
| **Median age (IQR), y** | 11.3 (5.3–15.8) | 11.4 (5.8–15.9) | 12.3 (7.3–16.8) |
| **Male sex, n (%)** | 17,111 (49.1) | 7,052 (47.6) | 832 (53.5) |
| **Ethnicity, n (%)** |  |  |  |
| Hispanic or Latino | 3,808 (11.0) | 1,421 (9.7) | 220 (14.2) |
| **Self-reported race, n (%)** |  |  |  |
| White | 21,975 (63.9) | 10,200 (69.6) | 887 (57.6) |
| Black | 5,218 (15.2) | 1,722 (11.8) | 339 (22.0) |
| Other | 4,733 (13.8) | 1,594 (10.9) | 219 (14.2) |
| Asian | 1,381 (4.0) | 582 (4.0) | 45 (2.9) |
| Multiple Races | 1,037 (3.0) | 520 (3.6) | 47 (3.1) |
| **Estimated median household income by zip code, \$** | 77,935 | 79,651 | 71,690 |
| **Social Vulnerability Index, n (%)** |  |  |  |
| Lowest | 15,019 (43.6) | 7,008 (47.4) | 551 (35.8) |
| Medium low | 9,013 (26.1) | 3,906 (26.4) | 395 (25.7) |
| Medium high | 5,306 (15.4) | 2,196 (14.8) | 256 (16.7) |
| Highest | 5,142 (14.9) | 1,679 (11.4) | 335 (21.8) |
| **New patient visits, n (%)** | 12,884 (37.1) | 3,429 (23.1) | - |

### Telemedicine use since the onset of the pandemic

Across the phases of the pandemic, we observed a dynamic pattern of telemedicine use (Figure 1). While >90% of all weekly new patient visits were performed by telemedicine during the shutdown phase (median 98%, interquartile range [IQR], 94%-99%) and ramp-up phase (median 96%, IQR, 84%-98%), the proportion of new patient visits via telemedicine decreased to 11% per week (IQR, 9%-19%) during the steady-state phase. Two outlier weeks can be observed during the steady-state phase (December 13, 2020 and January 31, 2021) during which most scheduled visits were converted to telemedicine due to snowstorms.

**Figure 1.**
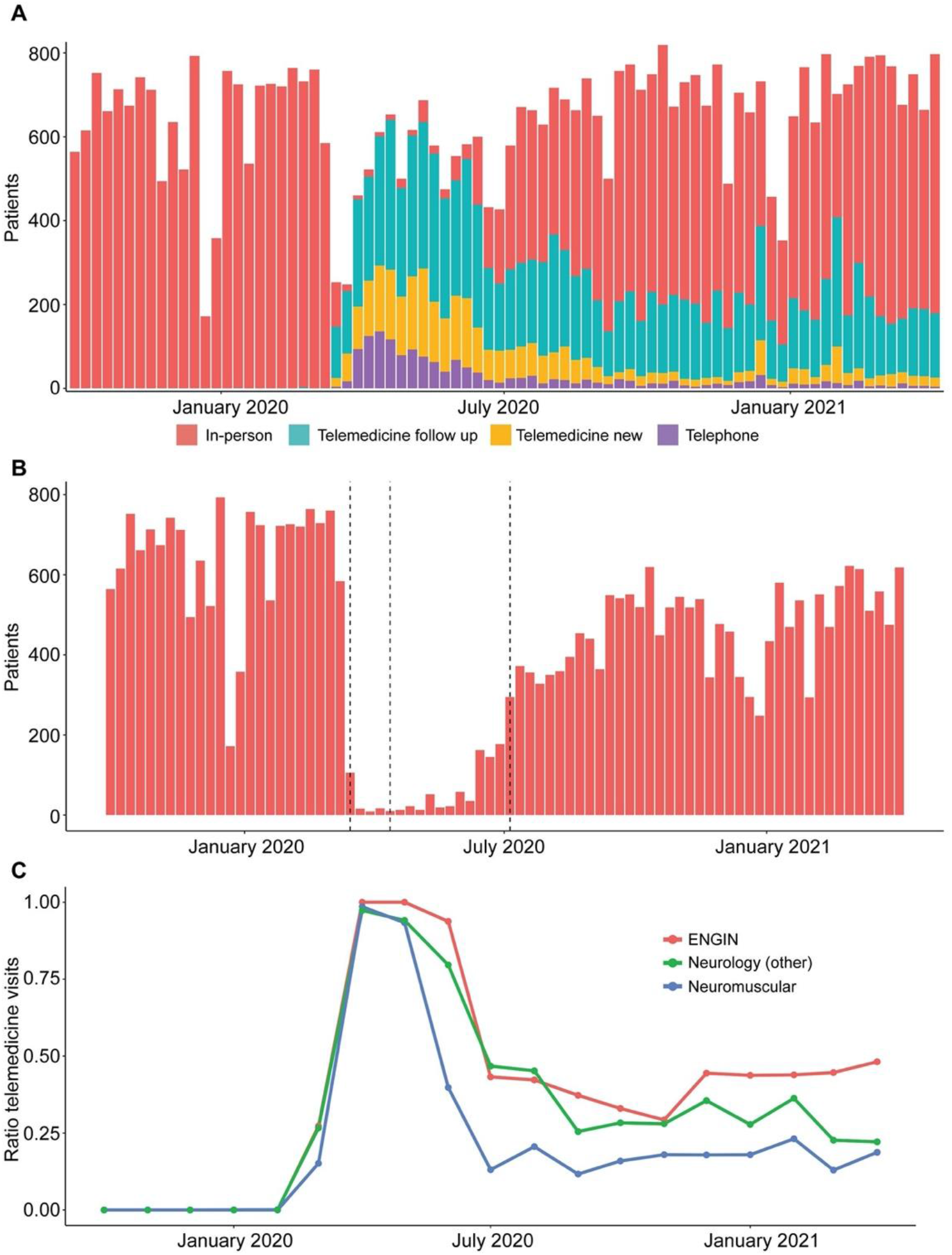
Child neurology outpatient encounters in response to the COVID-19 pandemic. (A) Volume of patient visits over time. (B) Volume of in-person visits during the phases of telemedicine use: prior phase, shutdown phase, ramp-up phase, steady-state phase. (C) Ratio of telemedicine visits across epilepsy neurogenetics (ENGIN), neuromuscular subspecialty program, and general child neurology clinic.

Of the 1,554 telephone visits performed during the entire observation period, there was an average of 75 (IQR, 17-125) weekly telephone visits during the shutdown phase, a much higher rate than the rest of the pandemic (OR 6.0, 95% CI 5.3 to 6.8). After this phase, telephone visits decreased but continued to be utilized at a low rate.

### Telemedicine in general child neurology and rare disease programs

We observed a differential effect of continued telemedicine use across broad community neurology programs and several specialty and subspecialty programs within our child neurology network (Figure 1C). We noted limited continued telemedicine use in the steady-state phase in general neurology clinics (28% of weekly visits, IQR, 25%-36%). In contrast, specific rare disease subspecialty programs either preferred in-person visits during this period, including less telemedicine use by the neuromuscular program (18% of weekly visits, IQR, 16%-19%), or continued telemedicine use by the epilepsy neurogenetics program (43% of weekly visits, IQR, 37%-44%).

### Telemedicine vs in-person visits

We compared diagnoses of individuals seen by telemedicine versus in-person visits (Figure 2A). Individuals with epilepsy (G40: OR 1.4, 95% CI 1.3-1.5) or attention deficit hyperactivity disorder (F90: OR 1.3, 95% CI 1.1-1.4) were more likely to receive care using telemedicine. Likewise, individuals with neurogenetic disorders including congenital syndromes (Q87: OR 1.4, 95% CI 1.1-1.6), microdeletions syndromes (Q93: OR 1.5, 95% CI 1.1-1.9) and neurogenetic disorders due to single gene mutations (Z15: OR 1.4, 95% CI 1.1-1.7) were more likely to receive care using telemedicine. In contrast, individuals with certain neuromuscular disorders were less likely to receive care by telemedicine, including spinal muscular atrophy (G12: OR 0.7 95% CI 0.5-0.9), muscular dystrophy (G71: OR 0.7, 95% CI 0.5-0.8) or other neuromuscular diagnoses (R29: OR 0.4, 95% CI 0.3-0.6; M62: OR 0.5, 95% CI 0.4-0.6). Similarly, individuals with movement disorders (G25: OR 0.7, 95% CI 0.6-0.8; R25: OR 0.7, 95% CI 0.6-0.9) or altered awareness (R40: OR 0.7, 95% CI 0.6-0.8) were less likely to receive care by telemedicine (Figure 2B). As some procedures and exams were put on hold during the first few months of the pandemic, there were specific diagnoses which were over-represented in the in-person visits during the ramp-up phase compared to in-person encounters prior to the shut-down or during the steady-state phase. These included individuals with migraine (G43) when comparing in-person visits in the ramp-up cohort to either the prior (OR 1.5, 95% CI 1.2-1.8) or the steady-state (OR 1.8, 95% CI 1.5-2.2) cohorts as well for individuals with spinal muscular atrophy (G12) or neuropathies (G60) during the ramp-up phase than during the prior phase (G12: OR 2.8, 95% CI 1.5-4.8; G60: OR 3.6, 95% CI 1.1-9.4) or during the steady-state phase (G12: OR 3.1, 95% CI 1.7-5.3; G60: 2.6, 95% CI 0.8-6.5). This suggests that a substantial proportion of individuals with these diagnoses may have received delayed care during the early phases of the pandemic and returned to in-person care after the initial shutdown in higher numbers compared to individuals with other diagnoses.

**Figure 2.**
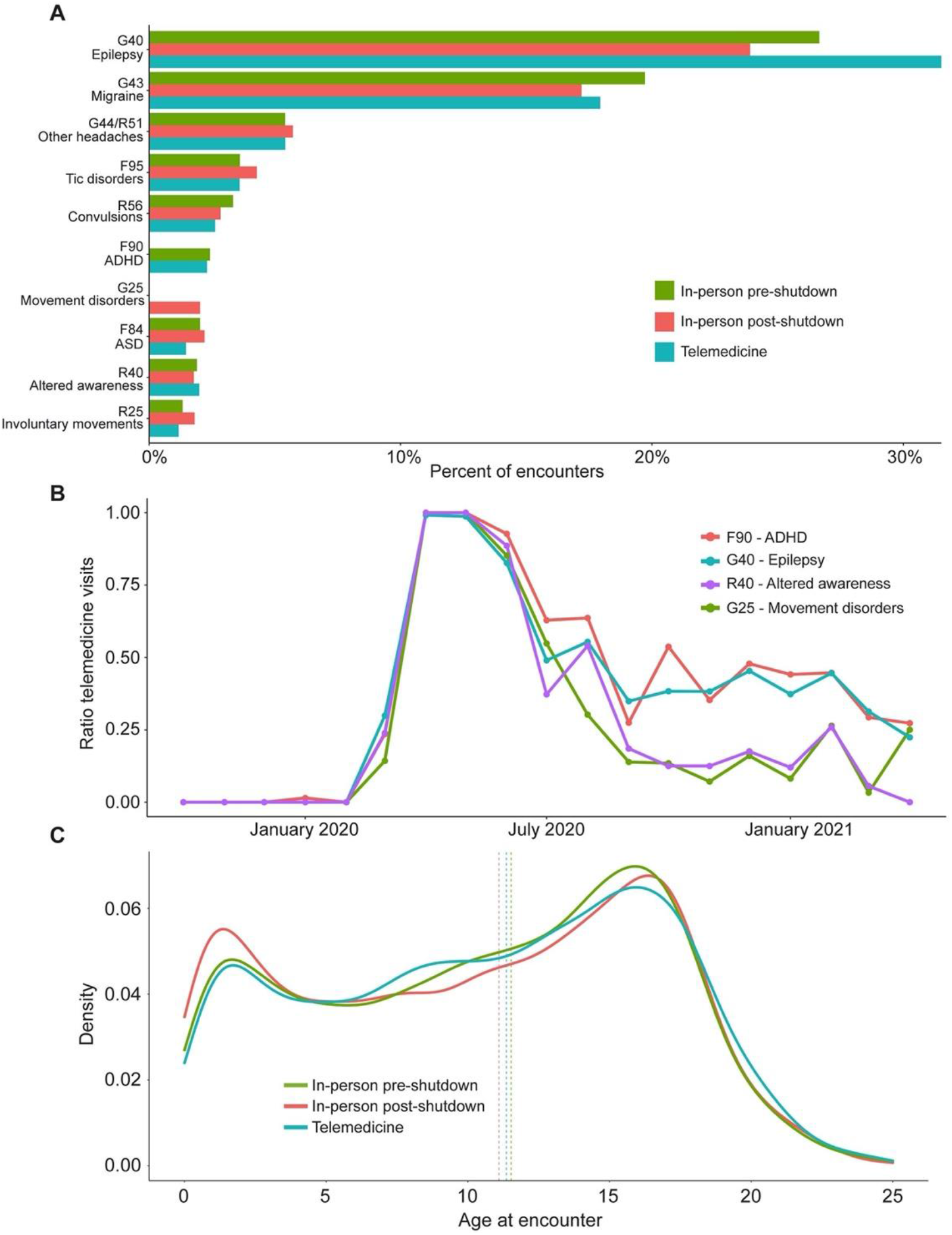
Diagnostic and age distribution of patients seen in-person and through telemedicine. (A) Spectrum of diagnoses in-person before and after transition to telemedicine. (B) Ratio of telemedicine visits within different diagnostic groups, highlighting the highest and lowest proportions of telemedicine use. (C) Age distribution of patients seen in-person pre- and post-shutdown and through telemedicine.

While the age distribution between the prior, shutdown, ramp-up, and steady-state cohorts were largely comparable, provider and family preferences likely led to some discrepancies. Infants below the age of one year were over 50% more likely to be seen in-person during all periods after the shutdown phase (6% of all visits during this period, OR 1.5, 95% CI 1.4-1.7) compared to all other phases and visit types (Figure 2C). Similarly, when compared against other age groups, infants were more likely to be seen in-person rather than via telemedicine during the shutdown phase (OR 2.1, 95% CI 1.3-3.7), and they accounted for 5% of all in-person visits during the shutdown.

### Equity of telemedicine access

We established two novel metrics to assess patient access to telemedicine (Figure 3). The first was the Patient Portal Activation Rate. Over the course of the pandemic, weekly activation rates rose from 75% to 86%. When assessing activation trends over the course of the pandemic, this rapid increase in activation occurred mostly during the first month of the pandemic (Kendall T=0.27, *P* <.01). By the end of the shutdown phase, the rise in activation stabilized (Kendall T=0.12, *P* =.22). There was a 13% difference in activation rates between highest and lowest SVI categories (OR 0.5, 95% CI 0.4-0.5). Among 15,823 visits in which individuals did not activate the patient portal, 4% (559/15,823) were scheduled telephone follow-ups and 92% (14,480/15,823) were scheduled in-person follow-ups, however 47% (7,474/15,823) of these visits were not completed due to cancellation or no-show.

**Figure 3.**
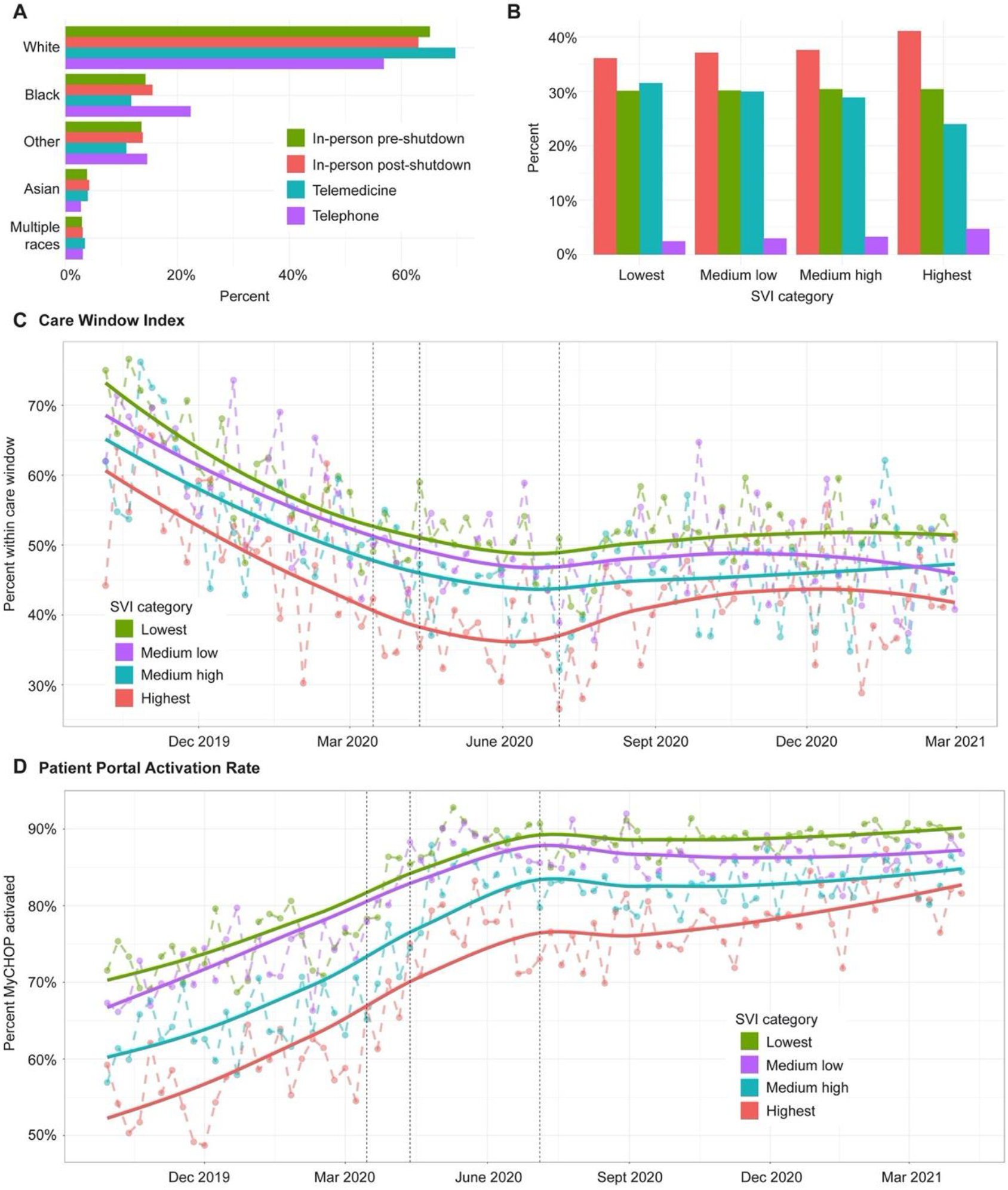
Follow-up within care window and patient portal activation rates as metrics to determine equity of telemedicine access. (A) Distribution of patient race in telemedicine, telephone and in-person visits. (B) Distribution of Social Vulnerability Index (SVI) categories in telemedicine, telephone and in-person visits. (C) Proportion of patients with follow-up within their care window over time. (D) Proportion of patients with successful portal activation over time.

Of the 2,705 individuals who were lost to follow-up, 423/2,705 (16%, OR 1.0, CI 95% CI 0.9-1.1) were in the highest SVI category. A total of 474/5,993 (8%, OR 2.6, CI 95% CI 2.3-2.9) individuals who did not activate the patient portal had a language other than English marked as their preferred language. Families in the highest SVI category (OR 1.8, 95% CI 1.5-2.1), with preferred language other than English (OR 2.0, 95% CI 1.5-2.6), or racial and ethnic minorities (OR 1.4, 95% CI 1.2-1.6) were more likely to have telephone visits.

The second novel metric was the CWI, indicating the proportion of individuals following up within an initially assigned time window. The average monthly ratio of individuals with delayed care (i.e., individuals not following up during the initially intended follow-up window) was 51% (SD ±9%). The percent of individuals with delayed care peaked early during the pandemic with up to 60% in July 2020 and remained high during the ramp-up and steady-state phases at 56% and 72% respectively. Even as individuals with specific diagnoses were overrepresented during certain phases, individuals with Tourette syndrome, convulsions, and headache were more likely to experience delayed care during the pandemic. Individuals in the highest SVI category (OR 1.4, 95% CI 1.3-1.5) and with a preferred language other than English (OR 1.2, 95% CI 1.0-1.3) were more likely to receive delayed care throughout all phases, particularly during the shutdown phase. Compared to individuals from less socially vulnerable families pre-pandemic, those in the highest SVI category were more likely to receive delayed care during the pandemic (OR 2.2, 95% CI 2.0-2.4).

## Discussion

The rapid adaptation of telemedicine in child neurology care due to the COVID-19 pandemic has been assessed in several studies,^13,15,20-25^ but beyond the initial implementation, the impact on care had not been fully assessed. Analyzing trends of telemedicine, a heterogeneous picture emerges with regards to patients suitable for telemedicine care and the effects of health disparities. Even when in-person visits became available, telemedicine continued to be used, albeit with variation across patient diagnoses, ages, and demographic factors.

The larger number of individuals and the longer evaluation period allowed us to identify patterns and perform comparisons that were previously difficult to assess. We identified that telemedicine is more frequent for individuals with epilepsy and attention deficit hyperactivity disorder, while in-person is likely preferred for individuals with migraines, spinal muscular atrophy, and neuropathies.

While ongoing management of pediatric epilepsies lends itself to telemedicine care, several populations are typically considered high risk for telemedicine evaluation, including young children and children with neuromuscular or movement disorders.^13-15^ We observed a lower rate of telemedicine visits in certain diagnosis groups and specialty programs with a noticeable increase in in-person visits during the ramp-up phase. Although we are unable to determine a unifying reason why a provider or family might prefer in-person visits, limitations such as the inability to complete certain procedures remotely and the ongoing pattern of preferred in-person evaluation emphasize the ongoing need to evaluate specific subgroups of individuals through in-person assessments.

We analyzed the effect of clinic re-opening, comparing telemedicine versus in-person assessments. When we compared cohorts in the ramp-up and steady-state phase, we were surprised to observe an increase in younger patients and patients with migraine in the in-person cohort. Previously, we had not observed this difference when comparing telemedicine during the shutdown versus prior in-person phases.^15^ We conclude that our previous comparison was incomplete and may have missed effects of delayed care or limited access. Accordingly, we estimate that initial assessments of telemedicine care early during the pandemic should be reassessed with a longer timeframe. Likewise, we postulate that adverse effects on specific patient groups caused by the widespread implementation of telemedicine may only become apparent once in-person care is accessible. This observation emphasizes the need to specifically target at-risk patient populations for follow-up during public health emergencies, such as the COVID-19 pandemic.

Underrepresented minorities and socially vulnerable families are at particular risk of receiving inadequate health care during public health emergencies. Limited access to telemedicine has emerged as a major factor in health disparities.^16,26^ We assessed whether specific obstacles in obtaining equitable care can be identified. To this end, we established two novel metrics to capture the effect of health disparities: a measurement to assess delayed care and a measurement to determine barriers to access telemedicine care once appointments are scheduled. We found that care across all groups was delayed by more than 50% during the onset of the pandemic and has remained delayed compared to pre-pandemic periods. In addition, individuals from highly socially vulnerable families had more than a two-fold risk for missing their follow-up care window during all phases of the pandemic. This compound effect highlights how socially vulnerable families were affected more severely during the pandemic with 423 individuals of the highest SVI category lost to follow-up at the completion of our study. Current projects within our institutions focus on approaching these families to resume care.

In parallel to the effect of health disparities on timely follow-up, we saw a strong effect of social vulnerability on the ability to engage in telemedicine, even when encounters were scheduled. We found an 11% difference in patient portal activation rates between families with lowest and highest SVI that persisted late into the pandemic, suggesting that many socially vulnerable families were not able to participate in telemedicine care. In our patient population, the lack of telemedicine care for more than 1,000 individuals can be attributed to social vulnerability.

## Limitations

Although this longer timeframe allowed us to make more comprehensive comparisons of telemedicine and in-person visits, certain variables remain beyond our current understanding. While we were able to determine how likely certain groups were to receive telemedicine versus in-person care, we could not measure this in relation to clinical outcomes. Although our healthcare network is currently in the process of integrating novel ways to assess clinical outcomes into our workflow in specific specialties,^27^ this is not yet widespread and records of varying aspects of clinical outcomes are incomplete.

Additionally, scheduling may have influenced the proportionality of certain visit types at different times during the pandemic, as certain procedures and exams had to be delayed until in-person care was available. This was especially true for families with infants, who were encouraged to be seen in-person, when possible, during the shutdown phase of the pandemic. As telemedicine becomes more integrated into neurology outpatient care, we plan to reassess whether these patterns remain.

Lastly, while we were able to analyze patient demographics metrics such as SVI, additional measurements could illuminate health disparities observed between groups. For example, integrating insurance information could further explain why certain families opted for in-person or telephone visits as opposed to telemedicine visits. Furthermore, comprehensive geographical analyses could help pinpoint regions where individuals may require additional outreach to prevent delay or loss to follow-up.

## Conclusion

Over the last year, telemedicine has emerged as a firm component of child neurology care. A substantial proportion of epilepsy care, especially for established patients, can be provided effectively by telemedicine. In contrast, care for individuals with neuromuscular conditions, movement disorders, and neurological assessment of young children typically require in-person assessments. Persistent telemedicine disparities emphasize the need for ongoing access to in-person visits during public health emergencies and the need to optimize telecommunications access for socially vulnerable families.

## Supporting information

EQUATOR STROBE Checklist

Disclosure form

## Data Availability

Data in a de-identified format will be made available by reasonable request to the
corresponding author.

## Acknowledgements

We thank the Arcus team at Children’s Hospital of Philadelphia for support with data analysis.

